# Seroprevalence of SARS-CoV-2 infection in the craft and manual worker population of Qatar

**DOI:** 10.1101/2020.11.24.20237719

**Authors:** Mohamed H. Al-Thani, Elmoubasher Farag, Roberto Bertollini, Hamad Eid Al Romaihi, Sami Abdeen, Ashraf Abdelkarim, Faisal Daraan, Ahmed Ismail, Nahid Mostafa, Mohamed Sahl, Jinan Suliman, Elias Tayar, Hasan Ali Kasem, Meynard J. A. Agsalog, Bassam K. Akkarathodiyil, Ayat A. Alkhalaf, Mohamed Morhaf M. H. Alakshar, Abdulsalam Ali A. H. Al-Qahtani, Monther H. A. Al-Shedifat, Anas Ansari, Ahmad Ali Ataalla, Sandeep Chougule, Abhilash K. K. V. Gopinathan, Feroz J. Poolakundan, Sanjay U. Ranbhise, Saed M. A. Saefan, Mohamed M. Thaivalappil, Abubacker S. Thoyalil, Inayath M. Umar, Zaina Al Kanaani, Abdullatif Al Khal, Einas Al Kuwari, Adeel A. Butt, Peter Coyle, Andrew Jeremijenko, Anvar Hassan Kaleeckal, Ali Nizar Latif, Riyazuddin Mohammad Shaik, Hanan F. Abdul Rahim, Hadi M. Yassine, Gheyath K. Nasrallah, Mohamed G. Al Kuwari, Odette Chaghoury, Hiam Chemaitelly, Laith J. Abu-Raddad, the Craft and Manual Workers Seroprevalence Study Group

**Affiliations:** Ministry of Public Health, Doha, Qatar; Hamad Medical Corporation, Doha, Qatar; Qatar Red Crescent Society, Doha, Qatar; College of Health Sciences, QU Health, Qatar University, Doha, Qatar; Biomedical Research Center, Qatar University, Doha, Qatar; Department of Biomedical Science, College of Health Sciences, Member of QU Health, Qatar University, Doha, Qatar; Primary Health Care Corporation, Doha, Qatar; Weill Cornell Medicine-Qatar, Cornell University, Doha, Qatar; Infectious Disease Epidemiology Group, Weill Cornell Medicine-Qatar, Cornell University, Doha, Qatar; World Health Organization Collaborating Centre for Disease Epidemiology Analytics on HIV/AIDS, Sexually Transmitted Infections, and Viral Hepatitis, Weill Cornell Medicine–Qatar, Cornell University, Qatar Foundation – Education City, Doha, Qatar; Department of Population Health Sciences, Weill Cornell Medicine, Cornell University, New York, New York, USA

**Author notes:** Address reprints requests or correspondence to Professor Laith J. Abu-Raddad, Infectious Disease Epidemiology Group, World Health Organization Collaborating Centre for Disease Epidemiology Analytics on HIV/AIDS, Sexually Transmitted Infections, and Viral Hepatitis, Weill Cornell Medicine - Qatar, Qatar Foundation - Education City, P.O. Box 24144, Doha, Qatar. **Disclose funding received for this work:** others.

**Keywords:** SARS-CoV-2, epidemiology, COVID-19, infection, seroprevalence, immunity, Qatar

## Abstract

**Background:** Qatar experienced a severe acute respiratory syndrome coronavirus 2 (SARS-CoV-2) epidemic that disproportionately affected the craft and manual worker (CMW) population who comprise 60% of the total population. This study aimed to assess the proportions of ever and/or current infection in this population.

**Methods:** A cross-sectional population-based survey was conducted during July 26-September 09, 2020 to assess both anti-SARS-CoV-2 positivity through serological testing and polymerase chain reaction (PCR) positivity through PCR testing. Associations with antibody and PCR positivity were identified through regression analyses.

**Results:** Study included 2,641 participants, 69.3% of whom were <40 years of age. Anti-SARS-CoV-2 positivity was estimated at 55.3% (95% CI: 53.3-57.3%) and was significantly associated with nationality, geographic location, educational attainment, occupation, presence of symptoms in the two weeks preceding the survey, and previous infection diagnosis. PCR positivity was assessed at 11.3% (95% CI: 9.9-12.8%) and was significantly associated with geographic location, contact with an infected person, and reporting two or more symptoms. Infection positivity (antibody and/or PCR positive) was assessed at 60.6% (95% CI: 9.9-12.8%). The proportion of antibody-positive CMWs that had a prior SARS-CoV-2 diagnosis was 9.3% (95% CI: 7.9-11.0%). Only seven infections were ever severe and one was ever critical—an infection severity rate of 0.5% (95% CI: 0.2-1.0%).

**Conclusions:** Six in every 10 CMWs have been infected, suggestive of reaching the herd immunity threshold. Infection severity was low with only one in every 200 infections progressing to be severe or critical. Only one in every 10 infections had been previously diagnosed suggestive of mostly asymptomatic or minimally mild infections.

## Introduction

The severe acute respiratory syndrome coronavirus 2 (SARS-CoV-2) infection has spread worldwide causing disease and mortality, as well as social and economic disruptions [1-3]. Qatar, a country in the Arabian Gulf, has experienced a pervasive epidemic with >55,000 laboratory-confirmed infections per million population as of November 20, 2020 [4, 5].

Most affected by the epidemic were the expatriate craft and manual workers (CMWs) who comprise 60% of the population of Qatar [6]. These workers are typically single men aged 20-49 years, recruited to work in development projects, and living in large shared accommodations [6-9]. Epidemiologic data on this population have indicated large SARS-CoV-2 outbreaks [7, 10, 11] that resembled those in nursing homes [12-14], or influenza outbreaks in regular and boarding schools [15, 16].

This study aimed to assess ever and current infection with SARS-CoV-2, infection severity rate, and infection diagnosis (detection) rate in the wider CMW population of Qatar.

## Methods

### Study design and sampling

A national cross-sectional survey was conducted between July 26 and September 09, 2020 to assess anti-SARS-CoV-2 (antibody) positivity and SARS-CoV-2 polymerase chain reaction (PCR) positivity among CMWs in Qatar. To optimize sample representativeness of the wider CMW population in the absence of a comprehensive listing for CMWs, we devised a sampling strategy based on analysis of the registered users’ database of the Qatar Red Crescent Society (QRCS), the main provider of primary healthcare for CMWs in the country.

The strategy applied sampling probability proportional to the size of registrants by age and nationality at each of the four QRCS health centers. The centers are distributed geographically factoring the distribution of CMWs in Qatar to cater to this population across the country. Sex was not considered as a variable in the sampling strategy because the vast majority of CMWs (>99%) are men [6]. The probability distribution of CMWs by age and nationality was cross-checked and found similar to that of the Ministry of Interior database of expatriate residents [8].

The overall sample size was determined at 2,232 assuming a seroprevalence of 25% (given the large epidemic in Qatar [7, 11]), a margin of error of 2%, and a non-response rate of 15%, but was increased to 2,658 to ensure that a minimum of five individuals were recruited per each age-nationality stratum from each center (for better representation of small groups).

Due to time constraints and operational challenges in contacting directly the CMWs and recruiting them, recruitment was implemented per the above sampling strategy but using systematic sampling of the attendees at these centers during the study duration. By factoring the average number of attendees per day at each of these centers, every 4^th^ attendee visiting each center was invited to participate in this study until the sample size by age and nationality at each center has been fulfilled. A written informed consent was collected from all study participants. The study was approved by Hamad Medical Corporation (HMC) and Weill Cornell Medicine-Qatar Institutional Review Boards.

### Sample collection and handling

An interview schedule inquiring about socio-demographics and history of exposure and symptoms was administered by trained interviewers in the participant’s language of preference. Both informed consent and interview schedule were provided and collected in nine languages (Arabic, Bengali, English, Hindi, Nepali, Sinhala, Tagalog, Tamil, and Urdu) to cater to the main language groups of CMWs. The study instrument was based on a protocol for SARS-CoV-2 sero-epidemiological surveys developed by the World Health Organization (WHO) [17]. Blood (10 ml) was drawn for serological testing by certified nurses and stored in an ice box before being transported to HMC Central Laboratory for analysis. Nasopharyngeal and oropharyngeal swabs were also collected by the nurses to assess current infection. National guidelines and standard of care were applied to all identified PCR positive cases. No action was mandated by national guidelines to those found antibody positive.

### Laboratory methods

Testing for SARS-CoV-2-specific antibodies in the serological samples was performed using an electrochemiluminescence immunoassay, the Roche Elecsys^®^ Anti-SARS-CoV-2 (Roche, Switzerland). Results’ interpretation was per manufacturer’s instructions: reactive for optical density cutoff index ≥1.0 and non-reactive for cutoff index <1.0 [18].

PCR testing was performed on aliquots of Universal Transport Medium (UTM) used for nasopharyngeal swabs’ collection (Huachenyang Technology, China). Aliquots were: extracted on the QIAsymphony platform (QIAGEN, USA) and tested with real-time reverse-transcription PCR (RT-qPCR) using the TaqPath™ COVID-19 Combo Kit (Thermo Fisher Scientific, USA) on a ABI 7500 FAST (Thermo Fisher, USA); extracted using a custom protocol [19] on a Hamilton Microlab STAR (Hamilton, USA) and tested using the AccuPower SARS-CoV-2 Real-Time RT-PCR Kit (Bioneer, Korea) on a ABI 7500 FAST; or loaded directly to a Roche cobas® 6800 system and assayed with the cobas® SARS-CoV-2 Test (Roche, Switzerland). All laboratory testing was conducted at HMC Central Laboratory following standardized protocols.

### Statistical analysis

Frequency distributions were used to characterize study participants. Absence/presence of symptoms in the two weeks preceding the survey (no symptoms, one, and two or more symptoms) was defined using a composite index score derived by summing up the values for reported symptoms coded as “0” for absence and “1” for presence. Probability weights were applied to adjust for participants’ unequal selection using CMW population distribution by age group and nationality per QRCS registered-user database.

Associations with anti-SARS-CoV-2 positivity were explored using Chi-square test and univariable logistic regression analyses. Covariates with p-value ≤0.2 in the univariable regression analysis were included in the multivariable model. Covariates with p-value ≤0.05 in the multivariable analysis were considered as showing strong evidence for an association with the outcome. Odds ratios (ORs), adjusted ORs (AORs), 95% confidence intervals (CIs), and p-values were reported. Associations with PCR positivity were also explored following above-described methodology.

Antibody test results were subsequently linked to the national SARS-CoV-2 PCR testing and COVID-19 hospitalization and severity database which includes all PCR testing, hospitalization, and SARS-CoV-2 infection severity classification as per the WHO criteria [20] since the start of the epidemic. Relevant epidemiological measures such as prevalence of ever and/or current infection, infection severity rate, and infection diagnosis rate were derived.

## Results

The final study sample included 2,641 participants, with a median age of 35 years (range: 18-80 years; Table 1). Most participants were below 40 years of age (69.3%) and of Indian (29.2%), Bangladeshi (26.2%), or Nepalese (21.6%) origin, representative of the wider CMW population in Qatar [8]. More than 40% had intermediate or lower educational attainment, and another 40% attended high school or vocational training. Over half of the sample consisted of technical and construction workers such as carpenters, crane operators, electricians, foremen, maintenance/air conditioning/cable technicians, masons, mechanics, painters, pipe-fitters, plumbers, and welders, while 4.7% held higher professional positions such as architects, designers, engineers, operation managers, and supervisors.

**Table 1.** Characteristics of study participants and associations with anti-SARS-CoV-2 positivity.

| Characteristics | Tested | Anti-SARS-CoV-2 positive |  | Univariable regression analysis |  |  | Multivariable regression analysis |  |
| --- | --- | --- | --- | --- | --- | --- | --- | --- |
|  | N (%*) | N (%†) | p-value | OR‡ (95% CI‡) | p-value | F test p-value§ | AOR‡ (95% CI‡) | p-value‡ |
| Age (years) |  |  |  |  |  |  |  |  |
| <29 | 753 (27.5) | 392 (53.2) | 0.279 | 1.00 |  | 0.296 | -- | -- |
| 30-39 | 979 (41.8) | 553 (57.5) |  | 1.19 (0.98-1.45) | 0.086 |  | -- | -- |
| 40-49 | 553 (21.5) | 298 (54.4) |  | 1.05 (0.83-1.32) | 0.690 |  | -- | -- |
| 50-59 | 265 (7.5) | 142 (54.9) |  | 1.07 (0.80-1.44) | 0.657 |  | -- | -- |
| 60+ | 91 (1.7) | 42 (47.6) |  | 0.80 (0.50-1.29) | 0.356 |  | -- | -- |
| Nationality |  |  |  |  |  |  |  |  |
| All other nationalities¶ | 231 (7.5) | 87 (38.2) | <0.001 | 1.00 |  | <0.001 | 1.00 |  |
| Filipino | 103 (2.7) | 25 (19.6) |  | 0.39 (0.22-0.70) | 0.002 |  | 0.45 (0.25-0.80) | 0.007 |
| Sri Lankan | 146 (4.8) | 56 (35.5) |  | 0.89 (0.56-1.40) | 0.614 |  | 0.79 (0.48-1.30) | 0.353 |
| Egyptian | 92 (3.2) | 32 (36.2) |  | 0.92 (0.54-1.56) | 0.748 |  | 0.89 (0.51-1.55) | 0.672 |
| Pakistani | 138 (4.9) | 70 (51.9) |  | 1.75 (1.11-2.73) | 0.015 |  | 1.34 (0.82-2.17) | 0.242 |
| Indian | 726 (29.2) | 376 (52.7) |  | 1.80 (1.31-2.47) | <0.001 |  | 1.50 (1.05-2.13) | 0.024 |
| Nepalese | 570 (21.6) | 345 (59.2) |  | 2.34 (1.68-3.26) | <0.001 |  | 1.75 (1.21-2.52) | 0.003 |
| Bangladeshi | 635 (26.2) | 436 (70.1) |  | 3.79 (2.72-5.26) | <0.001 |  | 2.95 (2.04-4.26) | <0.001 |
| QRCS center (catchment area within Qatar) |  |  |  |  |  |  |  |  |
| Fereej Abdel Aziz (Doha-East) | 618 (23.4) | 257 (42.5) | <0.001 | 1.00 |  | <0.001 | 1.00 |  |
| Hemaila (South-West; “Industrial Area”) | 981 (42.1) | 554 (57.6) |  | 1.84 (1.50-2.26) | <0.001 |  | 1.36 (1.00-1.85) | 0.050 |
| Zekreet (North-West) | 238 (2.3) | 125 (52.2) |  | 1.48 (1.09-2.00) | 0.012 |  | 1.42 (1.02-1.99) | 0.037 |
| Mesaimeer (Doha-South) | 804 (32.2) | 491 (61.7) |  | 2.18 (1.76-2.71) | <0.001 |  | 1.76 (1.38-2.24) | <0.001 |
| Educational attainment |  |  |  |  |  |  |  |  |
| Primary or lower | 633 (23.6) | 377 (60.2) | <0.001 | 1.00 |  | <0.001 | 1.00 |  |
| Intermediate | 434 (17.2) | 278 (64.8) |  | 1.22 (0.94-1.58) | 0.143 |  | 1.31 (0.99-1.73) | 0.055 |
| Secondary/High school/Vocational | 1,102 (42.5) | 599 (55.2) |  | 0.82 (0.66-1.00) | 0.054 |  | 1.08 (0.86-1.36) | 0.486 |
| University | 371 (12.7) | 114 (32.3) |  | 0.32 (0.24-0.42) | <0.001 |  | 0.70 (0.49-0.99) | 0.042 |
| Missing | 101 (4.0) | 59 (59.7) |  | 0.98 (0.63-1.53) | 0.934 |  | 1.29 (0.81-2.05) | 0.288 |
| Occupation |  |  |  |  |  |  |  |  |
| Professional workers‡ | 137 (4.7) | 36 (27.8) | <0.001 | 1.00 |  | <0.001 | 1.00 |  |
| Food & beverage workers | 93 (3.1) | 26 (29.2) |  | 1.07 (0.57-2.02) | 0.827 |  | 0.71 (0.37-1.37) | 0.304 |
| Administration workers | 82 (2.9) | 25 (31.6) |  | 1.20 (0.63-2.28) | 0.573 |  | 0.94 (0.49-1.82) | 0.857 |
| Retail workers | 171 (6.5) | 67 (40.3) |  | 1.76 (1.05-2.94) | 0.030 |  | 1.09 (0.64-1.87) | 0.753 |
| Cleaning workers | 105 (3.9) | 55 (54.5) |  | 3.12 (1.77-5.49) | <0.001 |  | 1.80 (0.98-3.31) | 0.060 |
| Transport workers | 435 (16.1) | 227 (53.4) |  | 2.99 (1.91-4.68) | <0.001 |  | 1.81 (1.12-2.94) | 0.016 |
| Technical and construction workers** | 1,329 (52.0) | 862 (65.1) |  | 4.85 (3.19-7.38) | <0.001 |  | 2.60 (1.66-4.09) | <0.001 |
| Security workers | 61 (2.3) | 36 (60.1) |  | 3.92 (2.02-7.61) | <0.001 |  | 2.90 (1.35-6.22) | 0.006 |
|  | N (%)<sup>*</sup> | N (%)<sup>†</sup> | p-value | OR<sup>‡</sup> (95% CI<sup>‡</sup>) | p-value | F test p-value<sup>§</sup> | AOR<sup>‡</sup> (95% CI<sup>‡</sup>) | p-value<sup>‡</sup> |
| Other workers<sup>††</sup> | 178 (6.8) | 64 (38.5) |  | 1.63 (0.98-2.71) | 0.061 |  | 1.23 (0.71-2.13) | 0.454 |
| Missing | 50 (1.7) | 29 (57.5) |  | 3.52 (1.69-7.30) | 0.001 |  | 2.28 (1.05-4.91) | 0.036 |
| Contact with infected person |  |  |  |  |  |  |  |  |
| No | 2,370 (89.3) | 1,283 (55.2) | 0.029 | 1.00 |  | 0.034<sup>§</sup> | -- | -- |
| Yes | 208 (7.9) | 101 (50.9) |  | 0.84 (0.63-1.13) | 0.258 |  | -- | -- |
| Unknown/Missing | 63 (2.8) | 43 (70.1) |  | 1.90 (1.10-3.29) | 0.022 |  | -- | -- |
| Symptoms in the two weeks preceding the survey |  |  |  |  |  |  |  |  |
| No symptoms | 2,326 (88.0) | 1,251 (54.6) | 0.021 | 1.00 |  | 0.021 | 1.00 |  |
| One symptom | 173 (6.7) | 107 (65.9) |  | 1.61 (1.14-2.26) | 0.006 |  | 1.56 (1.09-2.24) | 0.016 |
| Two or more symptoms | 142 (5.5) | 69 (53.3) |  | 0.95 (0.67-1.36) | 0.777 |  | 0.99 (0.68-1.43) | 0.946 |
| Symptoms required medical attention |  |  |  |  |  |  |  |  |
| No | 2,594 (98.0) | 1,392 (54.8) | 0.003 | 1.00 |  | 0.007<sup>§</sup> | -- | -- |
| Yes | 25 (1.0) | 17 (68.1) |  | 1.76 (0.73-4.21) | 0.205 |  | -- | -- |
| Unknown/Missing | 22 (0.9) | 18 (88.2) |  | 6.14 (1.81-20.89) | 0.004 |  | -- | -- |
| Symptoms required hospitalization |  |  |  |  |  |  |  |  |
| No | 2,613 (98.8) | 1,405 (55.0) | 0.020 | 1.00 |  | 0.020<sup>§</sup> | -- | -- |
| Yes | 2 (0.1) | 2 (100.0) |  | Omitted |  |  | -- | -- |
| Unknown/Missing | 26 (1.1) | 20 (79.0) |  | 3.09 (1.20-7.98) | 0.020 |  | -- | -- |
| Previously diagnosed with infection |  |  |  |  |  |  |  |  |
| No | 1,826 (66.0) | 957 (53.6) | 0.011 | 1.00 |  | 0.012 | 1.00 |  |
| Yes | 65 (2.5) | 45 (70.5) |  | 2.06 (1.19-3.57) | 0.010 |  | 1.96 (1.13-3.42) | 0.017 |
| Unknown/Missing | 750 (31.4) | 425 (57.5) |  | 1.17 (0.98-1.40) | 0.080 |  | 1.04 (0.78-1.39) | 0.769 |

AOR, adjusted odds ratio; CI, confidence interval; OR, odds ratio; QRCS, Qatar Red Crescent Society.
<sup>\*</sup>Percentage of the total sample weighted by age, nationality, and QRCS center.
<sup>†</sup>Percentage of positive out of those tested weighted by age, nationality, and QRCS center.
<sup>‡</sup>Estimates weighted by age, nationality, and center.
<sup>§</sup>Covariates with p-value ≤0.2 in the univariable analysis (for non-missing categories) were included in the multivariable analysis.
<sup>¶</sup>Covariates with p-value ≤0.05 in the multivariable analysis were considered as showing strong evidence for an association with anti-SARS-CoV-2 positivity.
<sup>‡</sup>Includes all other nationalities of craft and manual workers residing in Qatar.
<sup>§</sup>Includes architects, designers, engineers, operation managers, and supervisors among other professions.
<sup>¶</sup>Includes carpenters, construction workers, crane operators, electricians, foremen, maintenance/air conditioning/cable technicians, masons, mechanics, painters, pipe-fitters, plumbers, and welders among other professions.
<sup>††</sup>Includes barbers, firefighters, gardeners, farmers, fishermen, and physical fitness trainers among other professions.

A total of 1,427 participants had detectable SARS-CoV-2 antibodies—a seropositivity of 55.3% (95% CI: 53.3-57.3%; Table 2). Seropositivity was independently associated with each of nationality, QRCS center (proxy of catchment area/geographic location), educational attainment, occupation, presence of symptoms in the two weeks preceding the survey, and previous infection diagnosis in the multivariable regression analysis (Table 1). Still, the differences in seropositivity were overall not considerable apart from those by nationality, occupation, and geographic location (QRCS center). Compared to all other nationalities, AOR was 0.45 (95% CI: 0.25-0.80) for Filipinos, 1.50 (95% CI: 1.05-2.13) for Indians, 1.75 (95% CI: 1.21-2.52) for Nepalese, and 2.95 (95% CI: 2.04-4.26) for Bangladeshis. Compared to professional workers, AOR was 1.81 (95% CI: 1.12-2.94) for transport workers, 2.60 (95% CI: 1.66-4.09) for technical and construction workers, and 2.90 (95% CI: 1.35-6.22) for security workers. No association was found for age, contact with an infected person, symptoms requiring medical attention, or symptoms requiring hospitalization.

**Table 2.** Associations with SARS-CoV-2 polymerase chain reaction (PCR) positivity.

| Characteristics | Tested | SARS-CoV-2 PCR positive |  | Univariable regression analysis |  | F test p-value<sup>‡</sup> | Multivariable regression analysis |  |
| --- | --- | --- | --- | --- | --- | --- | --- | --- |
|  | N (%*) | N (%<sup>†</sup>) | p-value | OR (95% CI) | p-value |  | AOR (95% CI) | p-value<sup>§</sup> |
| Age (years) |  |  |  |  |  |  |  |  |
| <29 | 634 (29.4) | 82 (13.0) | 0.292 | 1.00 |  | 0.310 | -- | -- |
| 30-39 | 780 (42.0) | 77 (10.0) |  | 0.75 (0.53-1.05) | 0.090 |  | -- | -- |
| 40-49 | 408 (20.0) | 47 (11.6) |  | 0.88 (0.59-1.30) | 0.525 |  | -- | -- |
| 50-59 | 200 (7.1) | 24 (12.1) |  | 0.92 (0.56-1.54) | 0.762 |  | -- | -- |
| 60+ | 70 (1.6) | 3 (5.1) |  | 0.36 (0.10-1.31) | 0.121 |  | -- | -- |
| Nationality |  |  |  |  |  |  |  |  |
| All other nationalities<sup>‡</sup> | 202 (8.4) | 20 (10.3) | 0.099 | 1.00 |  | 0.110 | 1.00 |  |
| Indian | 549 (27.9) | 49 (8.4) |  | 0.80 (0.45-1.40) | 0.428 |  | 0.82 (0.45-1.51) | 0.527 |
| Sri Lankan | 114 (4.7) | 12 (11.6) |  | 1.14 (0.53-2.47) | 0.737 |  | 1.03 (0.47-2.26) | 0.935 |
| Egyptian | 79 (3.5) | 8 (11.4) |  | 1.12 (0.46-2.70) | 0.806 |  | 1.11 (0.44-2.81) | 0.820 |
| Bangladeshi | 496 (25.9) | 55 (11.4) |  | 1.12 (0.65-1.95) | 0.682 |  | 1.26 (0.70-2.27) | 0.448 |
| Pakistani | 116 (5.2) | 17 (14.7) |  | 1.50 (0.73-3.06) | 0.265 |  | 1.75 (0.82-3.75) | 0.149 |
| Nepalese | 467 (22.2) | 59 (13.3) |  | 1.34 (0.77-2.33) | 0.296 |  | 1.78 (0.99-3.21) | 0.053 |
| Filipino | 69 (2.2) | 13 (20.5) |  | 2.25 (0.99-5.11) | 0.053 |  | 2.22 (0.87-5.64) | 0.095 |
| QRCS center (catchment area within Qatar) |  |  |  |  |  |  |  |  |
| Fereej Abdel Aziz (Doha-East) | 547 (26.2) | 85 (15.4) | <0.001 | 1.00 |  | <0.001 | 1.00 |  |
| Mesaimeer (Doha-South) | 535 (27.0) | 21 (4.0) |  | 0.23 (0.14-0.37) | <0.001 |  | 0.30 (0.18-0.50) | <0.001 |
| Zekreet (North-West) | 186 (2.3) | 16 (8.8) |  | 0.53 (0.30-0.94) | 0.029 |  | 0.59 (0.33-1.08) | 0.087 |
| Hemaila (South-West; “Industrial Area”) | 824 (44.6) | 111 (13.4) |  | 0.85 (0.62-1.16) | 0.310 |  | 1.05 (0.73-1.51) | 0.801 |
| Educational attainment |  |  |  |  |  |  |  |  |
| Primary or lower | 502 (23.5) | 60 (12.5) | <0.001 | 1.00 |  | 0.204 | -- | -- |
| Intermediate | 347 (17.3) | 45 (12.5) |  | 1.00 (0.65-1.54) | 0.991 |  | -- | -- |
| Secondary/High school/Vocational | 870 (42.4) | 93 (11.2) |  | 0.89 (0.62-1.27) | 0.509 |  | -- | -- |
| University | 293 (12.8) | 31 (9.6) |  | 0.75 (0.46-1.22) | 0.245 |  | -- | -- |
| Missing | 80 (4.0) | 4 (4.3) |  | 0.31 (0.11-0.90) | 0.031 |  | -- | -- |
| Occupation |  |  |  |  |  |  |  |  |
| Professional workers<sup>¶</sup> | 107 (4.7) | 15 (13.9) | <0.001 | 1.00 |  | <0.001 | 1.00 |  |
| Cleaning workers | 85 (4.0) | 9 (7.9) |  | 0.53 (0.20-1.41) | 0.202 |  | 0.54 (0.18-1.56) | 0.253 |
| Security workers | 53 (2.5) | 3 (6.4) |  | 0.42 (0.11-1.55) | 0.192 |  | 0.55 (0.14-2.20) | 0.402 |
| Technical and construction workers<sup>‡</sup> | 1,041 (51.4) | 86 (8.5) |  | 0.57 (0.31-1.07) | 0.081 |  | 0.58 (0.29-1.15) | 0.118 |
| Administration workers | 70 (3.2) | 8 (12.1) |  | 0.85 (0.33-2.20) | 0.734 |  | 0.74 (0.27-2.03) | 0.559 |
| Food & beverage workers | 73 (3.1) | 12 (14.9) |  | 1.08 (0.44-2.64) | 0.859 |  | 0.89 (0.36-2.25) | 0.812 |
| Transport workers | 319 (14.7) | 38 (12.7) |  | 0.90 (0.46-1.77) | 0.753 |  | 0.94 (0.46-1.94) | 0.877 |
| Retail workers | 145 (7.0) | 27 (19.5) |  | 1.49 (0.72-3.07) | 0.278 |  | 1.46 (0.69-3.08) | 0.319 |
|  | N (%<sup>*</sup>) | N (%<sup>†</sup>) | p-value | OR (95% CI) | p-value |  | AOR (95% CI) | p-value<sup>§</sup> |
| Other workers<sup>**</sup> | 161 (7.7) | 32 (19.9) |  | 1.53 (0.76-3.10) | 0.234 |  | 1.60 (0.77-3.33) | 0.212 |
| Missing | 38 (1.7) | 3 (9.0) |  | 0.61 (0.16-2.30) | 0.464 |  | 0.63 (0.15-2.69) | 0.531 |
| Contact with infected person |  |  |  |  |  |  |  |  |
| No | 1,909 (90.7) | 195 (10.5) | <0.001 | 1.00 |  | <0.001 | 1.00 |  |
| Yes | 125 (6.1) | 36 (27.8) |  | 3.30 (2.14-5.11) | <0.001 |  | 2.87 (1.84-4.48) | <0.001 |
| Unknown/Missing | 58 (3.2) | 2 (2.8) |  | 0.25 (0.06-1.04) | 0.056 |  | 0.17 (0.04-0.76) | 0.021 |
| Symptoms in the two weeks preceding the survey |  |  |  |  |  |  |  |  |
| Asymptomatic | 1,856 (88.4) | 189 (10.5) | <0.001 | 1.00 |  | <0.001 | 1.00 |  |
| One symptom | 125 (6.0) | 15 (11.9) |  | 1.16 (0.65-2.08) | 0.615 |  | 1.20 (0.65-2.23) | 0.563 |
| Two or more symptoms | 111 (5.5) | 29 (23.5) |  | 2.63 (1.63-4.24) | <0.001 |  | 2.42 (1.47-3.96) | <0.001 |
| Symptoms required medical attention |  |  |  |  |  |  |  |  |
| No | 2,056 (98.1) | 229 (11.3) | 0.992 | 1.00 |  | 0.993 | -- | -- |
| Yes | 19 (0.03) | 2 (12.1) |  | 1.09 (0.25-4.78) | 0.914 |  | -- | -- |
| Unknown/Missing | 17 (1.2) | 2 (11.7) |  | 1.04 (0.24-4.62) | 0.954 |  | -- | -- |
| Symptoms required hospitalization |  |  |  |  |  |  |  |  |
| No | 2,069 (98.8) | 228 (11.1) | 0.108 | 1.00 |  | 0.079<sup>‡</sup> | -- | -- |
| Yes | 1 (0.03) | 0 (0.0) |  | Omitted |  |  | -- | -- |
| Unknown/Missing | 22 (1.2) | 5 (23.7) |  | 2.48 (0.90-6.83) | 0.079 |  | -- | -- |
| Previously diagnosed with infection |  |  |  |  |  |  |  |  |
| No | 1,428 (65.0) | 135 (9.4) | 0.001 | 1.00 |  | 0.002<sup>‡</sup> | -- | -- |
| Yes | 26 (1.2) | 5 (17.6) |  | 2.06 (0.75-5.63) | 0.161 |  | -- | -- |
| Unknown/Missing | 638 (33.8) | 93 (14.6) |  | 1.65 (1.23-2.20) | 0.001 |  | -- | -- |

AOR, adjusted odds ratio; CI, confidence interval; OR, odds ratio; QRCS, Qatar Red Crescent Society.
<sup>\*</sup>Percentage of the total sample weighted by age, nationality, and QRCS center.
<sup>†</sup>Percentage of positive out of those tested weighted by age, nationality, and QRCS center.
<sup>‡</sup>Covariates with p-value ≤0.2 in the univariable analysis (for non-missing categories) were included in the multivariable analysis.
<sup>§</sup>Covariates with p-value ≤0.05 in the multivariable analysis were considered as showing strong evidence for an association with anti-SARS-CoV-2 positivity.
<sup>†</sup>Includes all other nationalities of craft and manual workers residing in Qatar.
<sup>‡</sup>Includes architects, designers, engineers, operation managers, and supervisors among other professions.
<sup>§</sup>Includes carpenters, construction workers, crane operators, electricians, foremen, maintenance/air conditioning/cable technicians, masons, mechanics, painters, pipe-fitters, plumbers, and welders among other professions.
<sup>\*\*</sup>Includes barbers, firefighters, gardeners, farmers, fishermen, physical fitness trainers among other professions.

A total of 2,092 CMWs consented to PCR testing, of whom 233 had a positive result—a PCR positivity of 11.3% (95% CI: 9.9-12.8%). PCR cycle threshold (Ct) values ranged from 15.0-38.9 with a median of 27.6 (Figure 1). Ct value was ≥30 in 41.6% of PCR positive CMWs suggesting no active infection [21, 22]. PCR positivity was independently associated with geographic location (QRCS center), contact with an infected person, and reporting two or more symptoms in the two weeks preceding the survey in the multivariable regression analysis, but no association was found for the other variables (Table 2).

**Figure 1.**
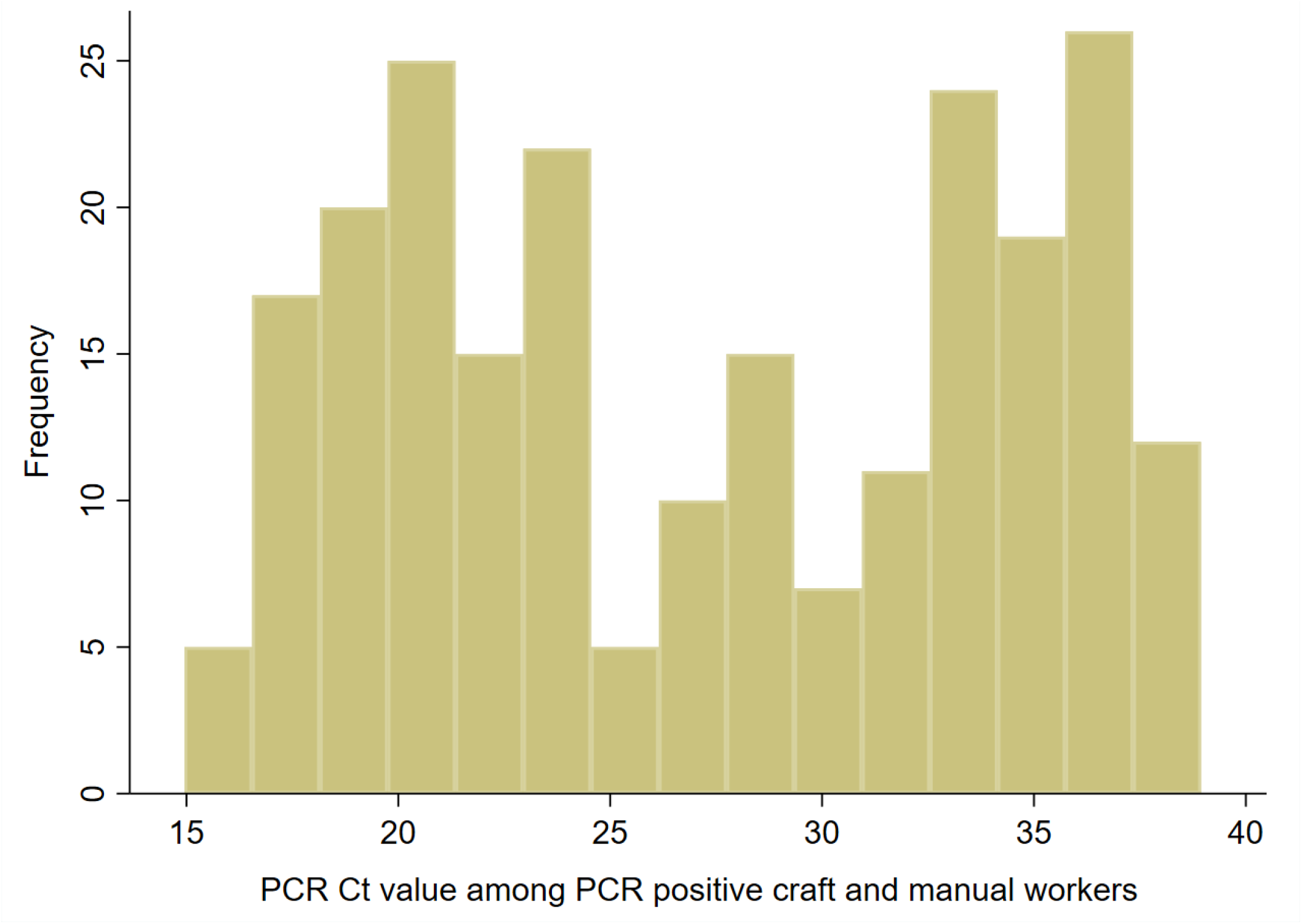
Distribution of polymerase chain reaction (PCR) cycle threshold (Ct) values among craft and manual workers (CMWs) identified as SARS-CoV-2 PCR positive during the study period.

Table 3 summarizes the key SARS-CoV-2 epidemiological measures assessed in this study. Infection positivity (antibody and/or PCR positive during the study) was assessed at 60.6% (95% CI: 9.9-12.8%). Of the 1,427 antibody-positive CMWs, 131 have had a laboratory-confirmed PCR positive result for SARS-CoV-2 *before* this study, corresponding to a diagnosis (detection) rate of 9.3% (95% CI: 7.9-11.0%). Median time between the previous PCR diagnosis and the antibody-positive test was 63 days. Meanwhile, 4 out of the 1,214 antibody-negative CMWs, 0.4% (95% CI: 0.1-1.0%), had been *previously* diagnosed with the infection prior to this study.

**Table 3.** Key SARS-CoV-2 epidemiological measures assessed in the study.

| **Epidemiological measure** | **Sample (denominator)** | **Positive for outcome (numerator)** | **Estimate (95% CI) in %\*** |
| --- | --- | --- | --- |
| Antibody positivity (seropositivity) prevalence | 2,641 | 1,427 | 55.3 (53.3-57.3) |
| PCR positivity prevalence | 2,092<sup>†</sup> | 233 | 11.3 (9.9-12.8) |
| Infection (antibody and/or PCR) positivity prevalence | 2,641 | 1,571 | 60.6 (58.6-62.5) |
| Infection diagnosis rate<sup>‡</sup> | 1,427 | 131 | 9.3 (7.9-11.0) |
| Antibody negative CMWs previously PCR-diagnosed with the infection | 1,214 | 4 | 0.4 (0.1-1.0) |
| Infection severity rate<sup>§</sup> | 1,590<sup>¶</sup> | 8<sup>¶</sup> | 0.5 (0.2-1.0) |

CMWs, craft and manual workers; PCR, polymerase chain reaction; CI, confidence interval.
\*Estimates weighted by age, nationality, and center.
<sup>†</sup>Only 2,092 persons consented to have a nasopharyngeal and oropharyngeal swab.
<sup>‡</sup>Proportion of antibody-positive CMWs with a prior SARS-CoV-2 laboratory-confirmed PCR diagnosis.
<sup>§</sup>Number of infections ever severe or critical per World Health Organization criteria over total number of laboratory-confirmed infections (antibody and/or PCR positive).
<sup>¶</sup>This number includes also 4 persons who were antibody negative and PCR negative at time of the survey, but had a PCR positive result prior to the survey. This number also includes 15 persons who were antibody negative and PCR negative at time of the survey, but had a PCR positive result subsequent to the survey at time of data linking and analysis (October 7, 2020).
\*Seven participants of this study have ever had (or progressed to) a severe infection and one had a critical infection per World Health Organization infection severity classification [20] at time of data linking and analysis (October 7, 2020).

Median time between the previous PCR diagnosis and the antibody-negative test was 28 days. The Ct values and PCR diagnosis date for these individuals were 16.0 on July 23, 22.3 on July 25, 22.8 on June 6, and 28.3 on May 2, 2020, suggesting that the recency of the infection may explain the lack of detectable antibodies for two of these four individuals.

Out of the total of 1,590 participants with laboratory-confirmed infection (antibody and/or PCR positive), seven have ever had or progressed to a severe infection (prior, during, or after this study) and one has ever had or progressed to a critical infection as per WHO criteria [20]—an infection severity rate of 0.5% (95% CI: 0.2-1.0%). All severe and critical infections have been hospitalized but cleared their infection; no COVID-19 deaths have been recorded.

## Discussion

The above results indicate that nearly two-thirds of the CMW population that constitutes 60% of the population of Qatar is at or near the herd immunity threshold for the SARS-CoV-2 infection.

This is to our knowledge the first such evidence for herd immunity in a majority segment of the population in any country. This conclusion is supported by the fact that no major infection cluster has been identified in any CMW community in Qatar for several months now, despite the progressive easing of the social and physical distancing restrictions since June 15, 2020 [23]. Meanwhile, large clusters of infection were common in such CMW communities before, around, and shortly after the epidemic peak towards the end of May, 2020.

The level of about 60-70% exposure to reach herd immunity is in concordance with that predicted using the “classical” formula for herd immunity of ◻1−1 *R*_0_ [24, 25], with *R0*, the basic reproduction number, being in the range of 2.5-4 [26, 27]. This, however, disagrees with assertions that herd immunity for SARS-CoV-2 infection can be reached at infection levels as low as 15-20% [28]. Our findings suggest that herd immunity may not be reached before at least half of the population has been infected, even in the presence of heterogeneity in the social contact rate in a given population [10, 25, 28].

A key finding of this study is the low SARS-CoV-2 infection severity rate found in this (relatively young) population where only one in every 200 infections was ever severe or critical as per the WHO infection severity classification [20]. This outcome agrees with findings of two other studies from Qatar where the infection severity rate has been estimated at 0.25% (95% CI: 0.11-0.49%) based on antibody and/or PCR laboratory-confirmed infections [10], and at 0.37% (95% CI: 0.37-0.38%) based on mathematical modeling of the epidemic in the total population [29]—compared to 0.50% (95% CI: 0.22-0.99%) in this study. These rates are substantially lower than those estimated elsewhere [30], often using early epidemic data, possibly because of insufficient accounting for the large denominator of undiagnosed asymptomatic or mild infections in young persons.

Notably, despite the large epidemic in Qatar, only 236 COVID-19 deaths have been registered as of November 21, 2020 [5], indicating also substantially lower infection fatality rate compared to earlier studies [30-33]. An analysis of the severity and fatality of SARS-CoV-2 infection in Qatar suggested the young age structure of the population, potential cross-reactivity to circulating ‘common cold’ coronaviruses, and high-quality standard of healthcare as reasons behind the low severity [29].

Though the infection was pervasive in this population, there were still some differences in exposure by nationality, catchment area/geographic location (QRCS center), educational attainment, and occupation (Tables 1 and 2). Given the totality of evidence on the Qatar epidemic [7, 10, 23, 29, 34], these differences may be explained by the nature of the shared accommodation (size and density), clustering of social networks by language and/or national background [7], occupational exposures (such as for drivers) [7], or differences in epidemic intensity in different parts of Qatar. Meanwhile, there were no differences in exposure by age.

The study had other notable findings. The study design allowed an empirical estimation of the diagnosis (detection) rate for this population. Out of all detected antibody-positive cases, only 9.3% (95% CI: 7.9-11.0%) had a documented PCR-confirmed infection *prior* to antibody testing in this study, indicating that nine in every 10 infections were never diagnosed, a finding that agrees with estimates from other settings [31, 35-38]. This outcome supports that most infections were asymptomatic or minimally mild to be diagnosed, in line with findings of a PCR community survey conducted earlier in Qatar in which 58.5% of those PCR positive reported no symptoms in the two weeks preceding the survey [7]. Another finding of the present study is that reporting of two or more symptoms was predictive of PCR positivity, but not reporting of only one symptom (Table 2), a similar finding to that of the earlier PCR community survey [7]. Lastly, a high proportion of those testing PCR positive had a Ct value >30, suggesting that nearly half of the PCR-positive CMWs may have acquired their infection 2-6 weeks earlier, given the common presence of prolonged PCR positivity in infected persons [21, 22].

This study had limitations. While the study design was intended to be based on probability-based sampling of the total CMW population in Qatar, operational challenges and time constraints forced instead a systematic sampling of QRCS attendees supplemented with probability-based weights to generate an estimate that is representative of the wider CMW population. It was difficult to recruit participants in the small age-nationality strata (such as among younger persons for specific nationalities), and thus towards the end of the study all attendees in these strata (not only every 4^th^ attendee) were approached to participate. Operational challenges made it also difficult to track and maintain consistent logs of the response rate by the nurses in these QRCS centers, thus an exact estimate of the response rate could not be ascertained, though it was estimated at >90% for antibody testing and at >70% for PCR testing. These limitations may have also introduced selection bias, such as biasing the assessed PCR positivity prevalence and infection severity rate towards higher values, as participants may have attended at the QRCS centers because of current-infection symptoms.

The laboratory methods were based on high-quality and validated commercial platforms, such as the Roche platform used for the serological testing [18, 39], one of the best available and extensively used and investigated commercial platforms with a specificity ≥99.8% [18, 40] and a sensitivity ≥95% [7, 39]. However, the less-than-perfect sensitivity may have lowered the measured antibody-positivity prevalence by a few percentage points.

In conclusion, six in every 10 CMWs have already been infected with SARS-CoV-2, indicating that this population is at or near herd immunity for this infection. While exposure to the infection was high, infection severity was low with only one in every 200 infections progressing to be severe or critical. Indeed, most infections must have been asymptomatic or too mild to be diagnosed, as only one in every 10 antibody-positive persons had a prior PCR-confirmed SARS-CoV-2 diagnosis.

## Data Availability

All data are available in aggregate form within the manuscript.

## Acknowledgements

We thank Her Excellency Dr. Hanan Al Kuwari, Minister of Public Health, for her vision, guidance, leadership, and support. We also thank Dr. Saad Al Kaabi, Chair of the System Wide Incident Command and Control (SWICC) Committee for the COVID-19 national healthcare response, for his leadership, analytical insights, and for his instrumental role in enacting data information systems that made these studies possible. We further extend our appreciation to the SWICC Committee and the Scientific Reference and Research Taskforce (SRRT) members for their informative input, scientific technical advice, and enriching discussions. We also thank Dr. Mariam Abdulmalik, CEO of the Primary Health Care Corporation and the Chairperson of the Tactical Community Command Group on COVID-19, as well as members of this committee, for providing support to the teams that worked on the field surveillance. We further thank Dr. Nahla Afifi, Director of Qatar Biobank (QBB), Ms. Tasneem Al-Hamad, Ms. Eiman Al-Khayat and the rest of the QBB team for their unwavering support in retrieving and analyzing samples and in compiling and generating databases for COVID-19 infection, as well as Dr. Asma Al-Thani, Chairperson of the Qatar Genome Programme Committee and Board Vice Chairperson of QBB, for her leadership of this effort. We also acknowledge the dedicated efforts of the Clinical Coding Team and the COVID-19 Mortality Review Team, both at Hamad Medical Corporation, and the Surveillance Team at the Ministry of Public Health. Last but not least, we thank all participants for their willingness to be part of this study.

## Author contributions

MHA and LJA co-conceived, co-designed, and co-led the study. EF led the study logistics and implementation. HC developed the study design, managed the databases, performed the data analyses, and wrote the first draft of the article. LJA led the statistical analyses and drafting of the article. All authors contributed to development of study protocol, data collection and acquisition, database development, discussion and interpretation of the results, and to the writing of the manuscript. All authors have read and approved the final manuscript.

## Craft and Manual Workers Seroprevalence Study Group

The Craft and Manual Workers Seroprevalence Study Group consists of personnel who have contributed to the implementation of this study.

### Qatar Red Crescent Society

Shafeer T. Aerattel, Firoj Ansari, Bennet J. Babu, Ali O. Bakari, Fazil K. Basheer, Muhammed J. Cherikkal, Muhammed R. Chonari, Ahmad S. Darwish, Arvin Dela Cruz, Verlili Z. Dela Cruz, Mark W. Del Carmen, Richie P. Deomampo, Sanu Gopi, Delfin JR O. Hortaleza, Robin Joseph, Veerankutty Kadar, Abdul Kareem A. Kalathil, Bigil C. Kandi, Mohammed M. T. Kaniyankandi, Kamarudheen Karimparukuzhiyil, Deelip G. Kurane, Manu Kurungott, Jommel R. C. Lumibao, Walid Mahmoud, Reyaz A. Malik, Jan A. Maxino, Nabeel T. Moosakutty, Hameed N. Nawabjahn, Ryan E. Orio, Mohamed F. Osman, Muhammad H. Ottappilakkool, Vijayakumar Pattakunninmel, Nissar P. Peedika, Suhail T. Puthiyaveettil, Ajith Raghavan, Renjee Ramachandran, Adil S. Sainudheen, Kannan Sassendran, John M. M. Soosai, Harris P. Sseri, Deepu Vallapil, and Patrick J. S. Venzuela.

### Ministry of Public Health

Rana A. M. Abdoon, Hind S. M. Ahmed, Ayah M. A. Mahmoud, Omnia O. E. Gismelkhalig, and Farid Shihata.

### Community Medicine Department at Hamad Medical Corporation

Khaled M. Ali and Fraih A. A. F. Alsallama.

## References

1. COVID-19 Outbreak Live Update. Available from: https://www.worldometers.info/coronavirus/. Accessed on September 06, 2020. 2020.

2. United Nations, Shared responsibility, global solidarity: Responding to the socio-economic impacts of COVID-19. Available from: https://www.un.org/sites/un2.un.org/files/sg_report_socio-economic_impact_of_covid19.pdf. Accessed on: April 16, 2020. 2020.

3. McKibbin W.J.F.R., The global macroeconomic impacts of COVID-19: seven scenarios. Available from: http://dx.doi.org/10.2139/ssrn.3547729 2020.

4. Hamad Medical Corporation, SARS-CoV-2 hospitalizations and care. 2020.

5. Ministry of Public Health-State of Qatar, Coronavirus Disease 2019 (COVID-19). Available from: https://covid19.moph.gov.qa/EN/Pages/default.aspx. Accessed on: May 25, 2020. 2020.

6. Planning and Statistics Authority-State of Qatar, Labor force sample survey. Available from: https://www.psa.gov.qa/en/statistics/Statistical%20Releases/Social/LaborForce/2017/statistical_analysis_labor_force_2017_En.pdf. Accessed on: May 01, 2020. 2017.

7. Abu-Raddad, L.J., et al., Characterizing the Qatar advanced-phase SARS-CoV-2 epidemic. medRxiv, 2020: p. 2020.07.16.20155317v2.

8. Ministry of Interior-State of Qatar, Population distribution by sex, age, and nationality: results of Kashef database. 2020.

9. De Bel-Air, F., Demography, Migration, and Labour Market in Qatar. Available from: https://www.researchgate.net/publication/323129801_Demography_Migration_and_Labour_Market_in_Qatar-_UPDATED_June_2017. Accessed on May 01, 2020. 2018, Gulf Labour Markets and Migration.

10. Jeremijenko, A., et al., Evidence for and level of herd immunity against SARS-CoV-2 infection: the ten-community study. medRxiv, 2020: p. 2020.09.24.20200543.

11. Al Kuwari, H.M., et al., Epidemiological investigation of the first 5685 cases of SARS-CoV-2 infection in Qatar, 28 February–18 April 2020. BMJ Open, 2020. 10(10): p. e040428.

12. Arons, M.M., et al., Presymptomatic SARS-CoV-2 Infections and Transmission in a Skilled Nursing Facility. N Engl J Med, 2020. 382(22): p. 2081–2090.

13. Burton, J.K., et al., Evolution and effects of COVID-19 outbreaks in care homes: a population analysis in 189 care homes in one geographical region of the UK. The Lancet Healthy Longevity, 2020. 1(1): p. e21–e31.

14. Ladhani, S.N., et al., Investigation of SARS-CoV-2 outbreaks in six care homes in London, April 2020. EClinicalMedicine, 2020. 26: p. 100533.

15. Jackson, C., et al., School closures and influenza: systematic review of epidemiological studies. BMJ Open, 2013. 3(2).

16. Glatman-Freedman, A., et al., Attack rates assessment of the 2009 pandemic H1N1 influenza A in children and their contacts: a systematic review and meta-analysis. PLoS One, 2012. 7(11): p. e50228.

17. World Health Organization, Population-based age-stratified seroepidemiological investigation protocol for COVID-19 virus infection. Available from: https://apps.who.int/iris/handle/10665/331656. Accessed on: April 15, 2020. 2020.

18. The Roche Group, Roche’s COVID-19 antibody test receives FDA Emergency Use Authorization and is available in markets accepting the CE mark. Available from: https://www.roche.com/media/releases/med-cor-2020-05-03.htm. Accessed on: June 5, 2020. 2020.

19. Kalikiri, M.K.R., et al., High-throughput extraction of SARS-CoV-2 RNA from nasopharyngeal swabs using solid-phase reverse immobilization beads. medRxiv, 2020: p. 2020.04.08.20055731.

20. World Health Organization, Clinical management of COVID-19. Available from: https://www.who.int/publications-detail/clinical-management-of-covid-19. Accessed on: May 31st 2020. 2020.

21. Sethuraman, N., S.S. Jeremiah, and A. Ryo, Interpreting Diagnostic Tests for SARS-CoV-2. JAMA, 2020.

22. Wajnberg, A., et al., Humoral immune response and prolonged PCR positivity in a cohort of 1343 SARS-CoV 2 patients in the New York City region. medRxiv, 2020: p. 2020.04.30.20085613.

23. Ayoub, H.H., et al., Mathematical modeling of the SARS-CoV-2 epidemic in Qatar and its impact on the national response to COVID-19. medRxiv, 2020: p. 2020.11.08.20184663.

24. Anderson, R.M., et al., How will country-based mitigation measures influence the course of the COVID-19 epidemic? Lancet, 2020. 395(10228): p. 931–934.

25. Britton, T., F. Ball, and P. Trapman, A mathematical model reveals the influence of population heterogeneity on herd immunity to SARS-CoV-2. Science, 2020. 369(6505): p. 846–849.

26. He, W., G.Y. Yi, and Y. Zhu, Estimation of the basic reproduction number, average incubation time, asymptomatic infection rate, and case fatality rate for COVID-19: Meta-analysis and sensitivity analysis. J Med Virol, 2020.

27. MIDAS Online COVID-19 Portal, COVID-19 parameter estimates: basic reproduction number. Available from: https://github.com/midas-network/COVID-19/tree/master/parameter_estimates/2019_novel_coronavirus. Accessed on: MAy 19, 2020. 2020.

28. Aguas, R., et al., Herd immunity thresholds for SARS-CoV-2 estimated from unfolding epidemics. medRxiv, 2020: p. 2020.07.23.20160762.

29. Seedat S. et al.., SARS-CoV-2 infection hospitalization, severity, criticality, and fatality rates in Qatar. under preparation, expected to be posted on medRxiv by Novemver 30, 2020.

30. Salje, H., et al., Estimating the burden of SARS-CoV-2 in France. Science, 2020.

31. Ioannidis, J.P., The infection fatality rate of COVID-19 inferred from seroprevalence data. Available at: https://www.medrxiv.org/content/10.1101/2020.05.13.20101253v1.full.pdf. Last accessed July 2, 2020. 2020.

32. Meyerowitz-Katz, G. and L. Merone, A systematic review and meta-analysis of Published research data on COVID-19 infection-fatality rates. Int J Infect Dis, 2020.

33. Hauser, A., et al., Estimation of SARS-CoV-2 mortality during the early stages of an epidemic: A modeling study in Hubei, China, and six regions in Europe. PLoS Med, 2020. 17(7): p. e1003189.

34. Abu-Raddad, L.J., et al., Assessment of the risk of SARS-CoV-2 reinfection in an intense re-exposure setting. medRxiv, 2020: p. 2020.08.24.20179457.

35. Anand, S., et al., Prevalence of SARS-CoV-2 antibodies in a large nationwide sample of patients on dialysis in the USA: a cross-sectional study. Lancet, 2020.

36. Havers, F.P., et al., Seroprevalence of Antibodies to SARS-CoV-2 in 10 Sites in the United States, March 23-May 12, 2020. JAMA Intern Med, 2020.

37. Wu, S.L., et al., Substantial underestimation of SARS-CoV-2 infection in the United States. Nat Commun, 2020. 11(1): p. 4507.

38. Stringhini, S., et al., Seroprevalence of anti-SARS-CoV-2 IgG antibodies in Geneva, Switzerland (SEROCoV-POP): a population-based study. Lancet, 2020. 396(10247): p. 313–319.

39. Jahrsdörfer, B., et al., Independent side-by-side validation and comparison of four serological platforms for SARS-CoV-2 antibody testing. The Journal of Infectious Diseases, 2020.

40. Public Health England, Evaluation of Roche Elecsys AntiSARS-CoV-2 serology assay for the detection of anti-SARS-CoV-2 antibodies. Available from: https://assets.publishing.service.gov.uk/government/uploads/system/uploads/attachment_data/file/891598/Evaluation_of_Roche_Elecsys_anti_SARS_CoV_2_PHE_200610_v8.1_FINAL.pdf. Accessed on June 5, 2020. 2020.

